# *Campylobacter* colonization and undernutrition in infants in rural Eastern Ethiopia: a longitudinal community-based birth cohort study

**DOI:** 10.1101/2024.05.21.24307707

**Authors:** Dehao Chen, Sarah Lindley McKune, Yang Yang, Ibsa Aliyi Usmane, Ibsa Abdusemed Ahmed, Jafer Kedir Amin, Abdulmuen Mohammed Ibrahim, Abadir Jemal Seran, Nurmohammad Shaik, Amanda Ojeda, Bahar Mummed Hassen, Loic Deblais, Belisa Usmael Ahmedo, Kedir Abdi Hassen, Mussie Bhrane, Xiaolong Li, Nitya Singh, Kedir Teji Roba, Nigel P. French, Gireesh Rajashekara, Mark J. Manary, Jemal Yusuf Hassen, Arie Hendrik Havelaar, the CAGED Research Team

## Abstract

**Background:** *Campylobacter* is associated with environmental enteric dysfunction (EED) and malnutrition in children. *Campylobacter* infection could be a critical link between determinants of livestock fecal exposure and health outcomes in low-resource smallholder settings.

**Methods:** We followed a birth cohort of 106 infants in a community of rural smallholder households in eastern Ethiopia up to 13 months of age. We measured anthropometry, surveyed socio-demographic determinants, and collected stool and urine samples. A short survey was conducted during monthly visits, infant stool samples were collected, and *Campylobacter* spp. was quantified using genus-specific qPCR. In month 13, we collected stool and urine samples to assay for biomarkers of EED. We employed regression analyses to assess the associations of household determinants with *Campylobacter* colonization, EED, and growth faltering.

**Results:** The *Campylobacter* load in infant stools increased with age. The mean length-for-age z-score (LAZ) decreased from −0.45 at 3-4 months of age to −2.06 at 13 months, while the prevalence of stunting increased from 3% to 51%. The prevalence of EED at 13 months of age was 56%. A higher *Campylobacter* load was associated with more frequent diarrhea. Prelacteal feeding significantly increased *Campylobacter* load in the first month of life. Over the whole follow-up period, *Campylobacter* load was increased by keeping chickens unconfined at home and unsanitary disposal of infant stools, while decreased by mother’s handwashing with soap. Longitudinally, *Campylobacter* load was *positively* associated with food insecurity, introduction of complementary foods, and raw milk consumption. There were no significant associations between *Campylobacter* load, EED, and LAZ.

**Conclusions:** This study found that most determinants associated with an increase in Campylobacter infection were related to suboptimal feeding practices and hygiene. Findings related to livestock-associated risks were inconclusive. Though stunting, EED, and *Campylobacter* prevalence rates all increased to *high* levels by the end of the first year of life, no significant association between them was identified. While additional research is needed to investigate whether findings from this study are replicated in other populations, community efforts to improve infant and young child feeding practices, including age at introduction of complementary foods and exclusive breastfeeding, and WaSH at the household level, could reduce (cross-) contamination at the point of exposure.

## Introduction

Stunting (length-for-age [LAZ]/height-for-age Z score <-2), affecting 22% of children under five (CU5) globally and 30% in sub-Saharan Africa [1], is an indicator of chronic undernutrition and is associated with numerous morbidities, all-cause mortality, and reduced lifetime earnings [2–4]. Agriculture-to-nutrition pathways, including household income, women’s empowerment, and agricultural production, can be leveraged to reduce undernutrition [5]. Particularly, animal source food (ASF) consumption has been associated with reducing child stunting in low- and middle-income countries (LMIC) [6].

Projected population and economic growth in Africa are expected to increase the demand for ASF fourfold between 2010 and 2050 [7]. Ethiopia is home to the largest livestock population on the continent, most of which is produced through extensive smallholder farming systems (i.e., mixed crop-livestock, pastoral) [8]. Resources are typically limited in such settings, and water, sanitation, and hygiene (WaSH) conditions often fail to meet international standards. This reality predisposes CU5 to environmental exposure to human and livestock feces, which are reservoirs of enteric pathogens [9].

Observational studies have associated enteric pathogens with child growth faltering [9,10], which is hypothesized to be mediated through acute illness (e.g., diarrhea, fever) [11] and environmental enteric dysfunction (EED), a chronic and asymptomatic consequence of both symptomatic and asymptomatic enteric infections [12]. EED is well established in literature as a risk factor for stunting [12–15], and growing evidence points toward EED as an important mediator between enteric infections and stunting [15]. The gold standard for diagnosing EED is intestinal biopsy [16], but it is not practical in population-based studies in resource-poor settings, where biomarkers were used to assess different impaired gut functions of EED [17]. However, a 2021 study from the SHINE trial found no consistent association between the examined EED biomarkers and linear growth [18]; and a 2022 study in two African countries revealed few correlations between the evaluated EED biomarkers [19]. Hence, challenges exist in the use of biomarkers to accurately measure EED.

As landmark randomized controlled trials focusing solely on reducing exposure to human feces through WaSH interventions found no significant effect on linear growth [20], researchers have called for a One Health approach that considers exposure to human and animal feces [21,22]. A systematic review of benefits and risks of smallholder livestock production found *Campylobacter* spp. to be the primary zoonotic enteric pathogen associated with increased risks of both EED and undernutrition and proposed a modification to the UNICEF framework on child undernutrition that incorporates poor gut health as a third immediate determinant of undernutrition [9]. These findings suggest that *Campylobacter* infection could be a critical link between livestock exposure and malnutrition in low-resource settings.

In 2018, the *Campylobacter* Genomics and Environmental Enteric Dysfunction (CAGED) research team conducted formative research, including a cross-sectional study of smallholder households in rural eastern Ethiopia [23]. This study found a high prevalence of infection with diverse *Campylobacter* spp., EED, and stunting in children aged 12-14 months and identified several associated risk factors [24]. These findings supported the design and implementation of a prospective longitudinal birth cohort study [25] to uncover the reservoirs and determinants of *Campylobacter* spp. colonizing infants, the results of which are presented here. Utilizing the modified UNICEF framework to identify possible putative determinants, this study examines associations in the first year of life between individual- and household-level determinants and *Campylobacter* infection. Given its importance to *Campylobacter,* we also explore symptoms of enteric illness (diarrhea and fever) as outcomes associated with *Campylobacter.* We then examine EED and linear growth faltering as secondary outcomes. Note that microbiological findings that detail the prevalence and load of Campylobacter from the CAGED longitudinal study have been described in a sister paper [26].

## Methods

### Study Setting

The CAGED longitudinal study was conducted in Haramaya woreda (district), East Hararghe Zone, Oromia Region, Ethiopia. The study site is a rural, agriculture-based area, where traditional crop-based agriculture has been replaced in recent years with cash crop production of khat; though many farmers still cultivate some corn and most have livestock. Data collection was scheduled to begin in April 2020 but was delayed due to the COVID-19 pandemic. After months of delay, preparation, and online training, data collection was initiated in December 2020, just one month after the start of the civil war in the Tigray Region, an armed conflict that continued throughout the data collection period [25].

### Study Design and Field Procedures

The study protocol has been described in full elsewhere [25]. Between December 2020 and June 2021, six monthly cohorts of up to 20 infants each, for a total of 115 infants, were randomly selected from a birth registry and enrolled in the study. Participating families were followed every four weeks (hereafter referred to as monthly follow-up) until the infants were approximately 13 months of age (through May 2022). We measured infant’s length and weight at quarterly visits following enrollment using the same procedures described in the formative research [24]. Despite extensive efforts to engage with culturally sensitive approaches, some mothers were reluctant to hand over newborn infants to local enumerators for measurement at birth. In addition, birth length measures are error-prone due to infants being bent and curled up [27]. For these reasons, we excluded length measures shortly after birth from our analyses.

A Long Survey with mothers and their male partners was conducted at baseline and near the end of the study to collect information including demographics; livelihoods; wealth; animal ownership, management, and diseases; WaSH conditions; and child health. During monthly visits, mothers responded to a Short Survey focusing on dynamic indicators, including infant health, diets, food security, contact with animals, and the environment; infant stool samples were collected concurrently. All interview, survey, and anthropometry data were collected on tablets using the REDCap mobile app and were uploaded to a secure REDCap database at the University of Florida. All survey instruments are available in a supplementary file to the study protocol manuscript [25].

At the end of the follow-up period, samples were collected to assess biomarkers of EED as previously reported [24]. Briefly, stool samples were collected to measure fecal myeloperoxidase (MPO) and immediately flash-frozen using liquid nitrogen and stored at −80 L until further use. To assess gut permeability, a solution of 1000 mg lactulose in 10 mL sterile water was administered to the infant for 5 min, and urine was collected over a period of 4 hours. All samples were transported to the USA on dry ice for further analysis. MPO was measured using a commercially available enzyme-linked immunosorbent assay (MPO RUO, Alpco, Salem, NH). Lactulose in urine was assessed using highLperformance liquid chromatography (HPLC).

### Sampling and Laboratory Procedures

The collection and preservation of infant stool samples and molecular detection and quantification of *Campylobacter* by quantitative polymerase chain reaction (qPCR) have been described elsewhere [26]. The primers employed detect a large range of species in the *Campylobacter* genus, including the well-studied species *Campylobacter jejuni* and *C. coli* as well as a wide range of other thermotolerant and non-thermotolerant species such as the recently described Candidatus *C. infans* [28]. Based on the standard curve of qPCR, *Campylobacter* load (unit: log_10_[gene copies per 50 ng of DNA]) was calculated as a linear function of cycle threshold (Ct) values: 10.51-0.24*Ct [26]. We chose not to use a Ct-cutoff value to categorize the presence/absence status of *Campylobacter* in the following data analyses to avoid left censoring of load estimates (i.e., false negative when the load is under a certain detection threshold [non-detectable]).

### Data Analyses

#### Health outcomes

The primary outcome evaluated in this study is the *Campylobacter* load of infants (including those with and without clinical symptoms). We categorized the ages when the infant stool samples were collected into quartiles to assess temporal variations of the *Campylobacter* load and its determinants.

Secondary outcomes include EED, linear growth faltering, diarrhea, and fever. EED was categorized using a composite indicator based on the percentage of lactulose excretion (%L) and fecal MPO [24]. Cut-off values for %L were 0.2 < %L ≤ 0.45% for moderate gut permeability and %L > 0.45 for severe gut permeability. The thresholds for MPO were 2,000 ng/ml for moderate gut inflammation and the third quartile of the observed data 3364 ng/ml) for severe gut inflammation. EED was defined as moderate if either %L or MPO was categorized as severe; and as severe if both %L *and* MPO were in a severe category. Linear growth faltering was characterized by a decrease in LAZ. We modeled changes in LAZ using two models: one uses the overall decrease between the first and last available LAZ scores as the outcome, referred to as the decrease in LAZ, and is a non-longitudinal model. The other is a longitudinal model, using the LAZ at each visit as the outcome. We use the phrases “linear growth faltering” and “decrease in LAZ” interchangeably. As the anthropometric measurements were conducted on different days than the short surveys and stool sample collection, we employed k-means clustering to categorize the ages at anthropometric measurements into 5 groups. Neonatal outcomes include the *Campylobacter* load in the first month of life (day range: [7–39]) and the first available LAZ. A case of diarrhea or fever was declared when the mother reported the infant having symptoms on the day of the interview or the day before. The definition of diarrhea provided to the mother was the infant having three or more loose or watery stools in 24 hours.

#### Identification of putative determinants

We applied the modified United Nations Children’s Fund (UNICEF) framework of child undernutrition, as proposed in our systematized review [9], to identify putative determinants of the health outcomes of interest (*Campylobacter* infection, EED, and linear growth faltering). These included variables from the household surveys that fell into domains of *immediate causes* (*inadequate dietary intake* and *diseases*), *underlying causes* (*household food insecurity*, *inadequate care and feeding practices*, and *unhealthy household environment and inadequate health services*), and *basic causes* (*social/cultural*, *economics/livelihood*, *human capital*, *basic demographics*) from the original UNICEF framework, as well as variables that reflected the domains of *benefits* (as determined by the agriculture-to-nutrition pathways), *risks* (of enteric infections from livestock exposure), and *control measures* (to the *risks*), as illustrated in the review. For the conceptual overlap between *risks* and *unhealthy household environment* underlying *Campylobacter* infection, we assigned the determinants related to livestock (feces) exposure to *risks*, given our study’s interest on smallholder farming. The determination of eligible variables for analyses involved cross-checking the framework’s domains and the variables in the household surveys, generating a suite of putative determinants for the health outcomes of *Campylobacter* infection, EED, and linear growth faltering. Some determinants may affect the down-streaming outcomes of poor gut health and undernutrition through other mediators, but may not be directly associated with an increased risk of *Campylobacter* infection. These determinants include vitamin A and iron supplementation, treatment for malnutrition, use of oral rehydration solution, visiting a health center for diarrhea or fever, vaccination, and receiving prenatal care and its location; thus, we only tested associations with EED and LAZ to simplify interpretation. We examined the enteric symptoms of diarrhea and fever as possible outcomes of *Campylobacter* infection. We also considered both enteric symptoms as determinants of growth faltering. While fever was considered a putative determinant for EED, diarrhea was not, as diarrhea is a causative factor for elevated EED biomarkers.

#### Composite variables

Composite variables were calculated by combining answers from two or more survey questions, and such variables have been validated in other studies. We generated the following composite variables:

1. Minimum Dietary Diversity of Infant and Young Children (MDD-IYC) [29], from which the consumption status of ASF was extracted.
2. Household food insecurity access score (HFIAS) and household food insecurity access (HFIA) category [30].
3. *Joint Monitoring Programme* (JMP) WaSH service ladders for drinking water, sanitation, and hygiene [31].
4. Tropical Livestock Units (TLU), a composite variable that quantifies the holding of all farmed animals in a household [32].
5. Anthropometric Z scores (i.e., LAZ and weight-for-age Z [WAZ] scores), calculated using the R package *anthro* [33].
6. Adapted from our formative research [24], we defined a livestock location risk score for each of four livestock species (cattle, chicken, sheep, or goat). This score was designed to characterize increasing likelihood of contact between a livestock species and study participants during the day or night. A score of 0 was assigned to any household that did not have the livestock species in question or reported keeping them outside the house at all times; a score of 1 was assigned to any household that reported keeping the livestock species inside the house and confined; and a score of 2 was assigned to any household that reported keeping the livestock species inside the house unconfined.
7. To characterize assets, informed by the literature [34], we employed a latent trait model (using the R package *ltm* [35]) to calculate a factor score for ownerships of sellable household assets for each household.
8. We characterized the age of introducing complementary feeding as the infant’s age at the monthly visit when the mother first reported giving the infant complementary foods. This definition of complementary feeding is based on operational definitions of WHO [36], which do not consider prelacteal feeding.
9. Complementary feeding refers to an infant receiving any solid, semisolid, or soft foods at any time (note that this differs from appropriate complementary feeding which should occur after six months of age).
10. Prelacteal feeding means feeding any substances other than breastmilk given to the infant during the first three days after birth.

To facilitate interpretation and minimize the impact of data sparsity, we dichotomized some composite variables. We used accepted cutoffs for meeting MDD as ≥ 5, a HFIA category of 1 as food secure, and an anthropometric Z score < −2 as being stunted. In the JMP drinking water, sanitation, and hygiene ladders, the categories of *unimproved*, *open defecation*, and *no facility* were treated as reference categories, respectively, with all other categories within each ladder merged. Infant stool disposal was binarized as a low-risk category including infant using toilet or latrine, stool put or rinsed into toilet or latrine or drain or ditch and a high-risk category including stool thrown in garbage, buried, or left in the open. The quartile score for assets was binarized by its median.

#### Regression analyses

The average *Campylobacter* load, EED, and the change in LAZ for each infant over the follow-up period were separately regressed on all selected baseline determinants, except for infant feeding practices at birth. As we were interested in the short-term effects of feeding practices at birth (colostrum feeding, early initiation of breastfeeding, and prelacteal feeding) on *Campylobacter* infection and on linear growth, we assessed their relationship with the *Campylobacter* load in month one (age range [7–39]) and the first available LAZ measurement in the regression analysis. EED at endline was regressed on all the selected baseline determinants and the average of longitudinal (time-varying) determinants of the whole follow-up.

To assess the longitudinal associations between the time-varying determinants and health outcomes, we regressed the average *Campylobacter* load on the average of each time-varying determinant within each age quartile, under the assumption that the effect of each determinant was immediate to the change of *Campylobacter* load. We used the same approach to regress separately the outcomes of current fever and diarrhea on *Campylobacter* load. In contrast, the LAZ of each visit was regressed on the average value of each determinant from birth up to that visit, under the assumption that stunting is the result of a chronic process and the effect of each determinant on growth accumulates over time.

We employed linear-mixed models with an individual-level random intercept to account for dependency in repeated measures (using the R package *lme4* [37]). For each outcome (except diarrhea and fever), we conducted backward elimination to select a multivariable model, starting with determinants having adjusted p values < 0.2 from a variable screening step where the outcome was regressed on each determinant separately while adjusting for sex and socio-economic status (SES, binarized household asset score) as fixed confounders. Infant age group was also adjusted as a fixed confounder in the linear-mixed models for *Campylobacter* load and LAZ, while the age at EED sampling was adjusted as a fixed confounder in the logistic models for EED. These fixed confounders were also constantly kept in the multivariable models, i.e., they are not subject to backward elimination in the multivariable selection. In the models for EED, we categorized variables of Proportion of Vitamin A supplementation (categories: ≥ 0.15, > 0 & < 0.15, and =0) and proportion of infants with fever (categories: ≥0.3, <0.3) to improve the interpretability of the associated effects and the readability of their confidence intervals. For most variables, there was a higher level of missingness at the baseline than at the endline. For variables that are the same between baseline and endline among >80% of the studied households, and if the baseline value was missing but the endline value was available, we used the endline value to impute the missing baseline value. After this simple ad hoc imputation, we conducted complete case analyses, i.e., missing data were dropped under the assumption of missing completely at random. All analyses were conducted in *R* (version 4.1.0) [38].

## Results

### Descriptive statistics

Of the 115 infants enrolled in the study, four households withdrew from the study, three infants died from causes unrelated to the study, and two moved outside of the study area, leaving 106 infants who completed follow-up and were included in the analysis. Of the 1,378 planned household visits, 1,111 (81%) had infant’s stool samples assayed by qPCR, 1,035 (75%) had a monthly short survey, and 980 (71%) had both. Only 13% and 2% of infants had stool samples collected and short surveys completed for all 13 visits, respectively. We present characteristics of the study population in Table 1.

**Table 1.**
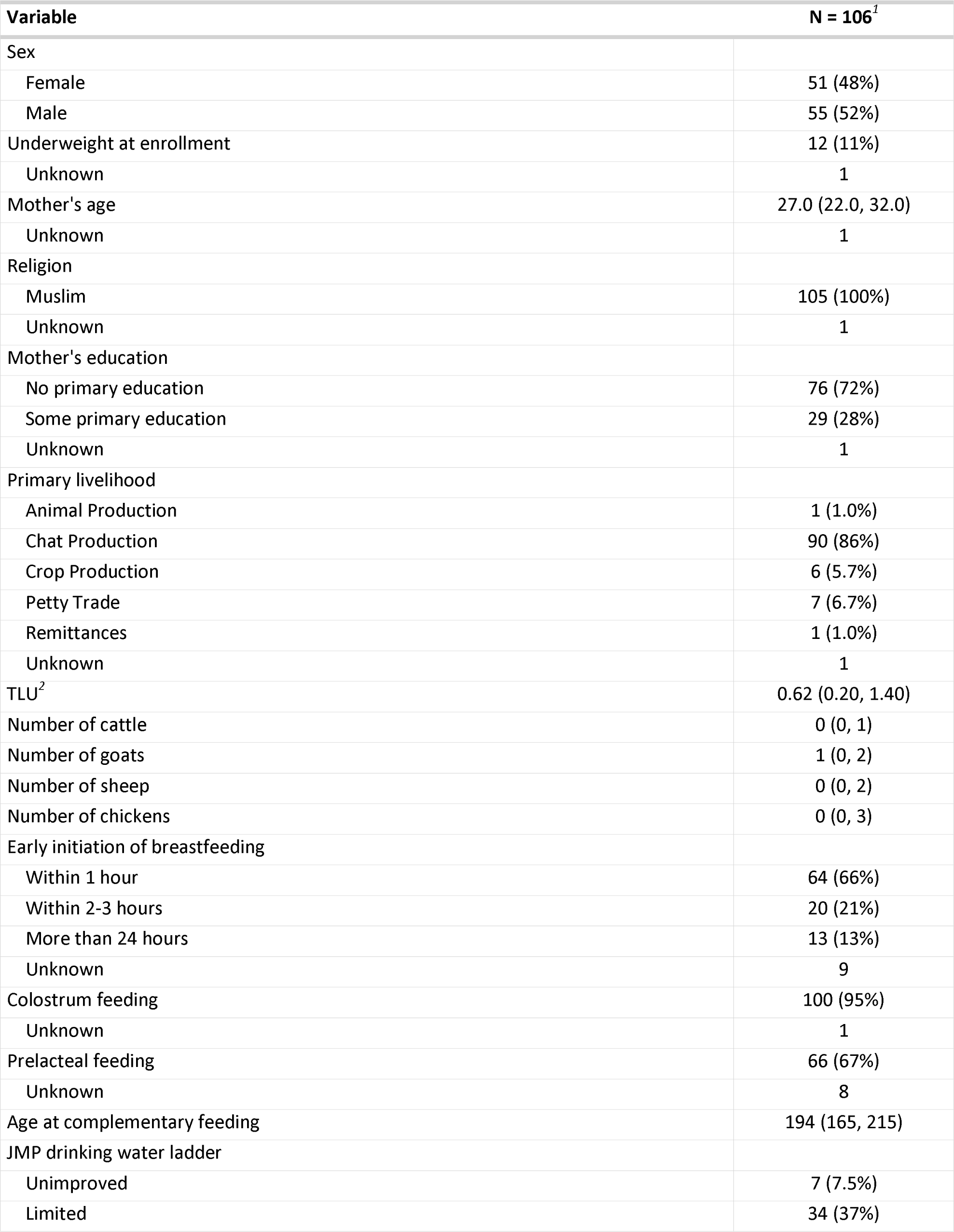
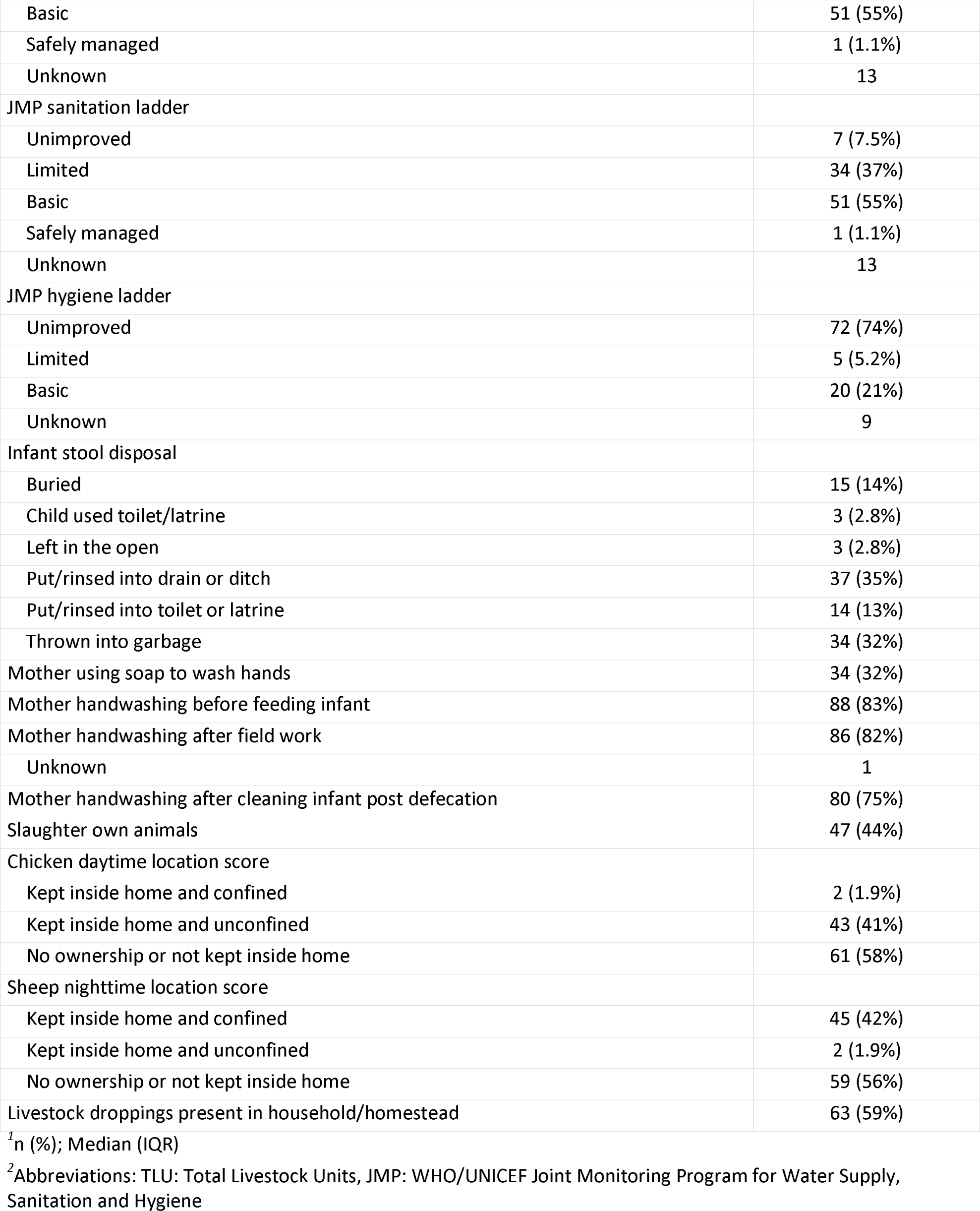
Characteristics of the study population.

Forty-eight percent of infants were females, and 11% were underweight at enrollment (age range: [4–27] days). The mothers were on average 27 years old, all of whom were Muslim, and 28% had received some formal education. Most households were engaged in a combination of on-farm activities (e.g., animal, crops and khat production), with khat production dominating as the primary livelihood for 86% of households and an average TLU of 0.62. Almost all infants were fed colostrum, and 66% had early initiation of breastfeeding (within one hour) after birth; 67% received prelacteal feeding. Few households had access to safely managed drinking water (1.1%) and sanitation facilities (1.1%), and only 21% of households had access to basic hygiene facilities. More than three quarters of mothers reported hand washing before feeding the infant, after fieldwork, and after infant defecation; however, soap use was only reported by one third of mothers. Most infant stools were reportedly disposed of as garbage or into a drain. During the daytime, all farm animals except chickens were kept either outside of the household or confined if inside. More farm animals (goats, sheep, cattle, and chickens) were kept inside at night than during the day. The proportion of households who confined chickens when kept inside the home (41%) was higher than for other livestock and was the same during the day- or nighttime. While 59% of households were observed to have livestock droppings in household/homestead, nearly 90% of households reported collecting animal waste. Forty-four percent of households reported slaughtering their own livestock (goat, sheep, or chicken), and all these households reported washing hands before and/or after slaughtering. Further details of household characteristics were reported in supplementary Table 1.

In the first month of life (day range: [7–39]), the average *Campylobacter* load in infant stools was 1.87 (SD: 1.01, unit: log[gene copies / 50 ng of DNA]), whose corresponding prevalence was 32% (95% CI: 24%-42%), under a Ct cutoff of 35. The *Campylobacter* load increased steadily from 1.95 (SD: 0.97) in the first age quartile to 3.83 (SD: 1.14) in the fourth age quartile, see Figure 1(A). The average load corresponding to each anthropometry visit also demonstrated a steadily increasing trend, see Figure 1 (B).

**Figure 1.**
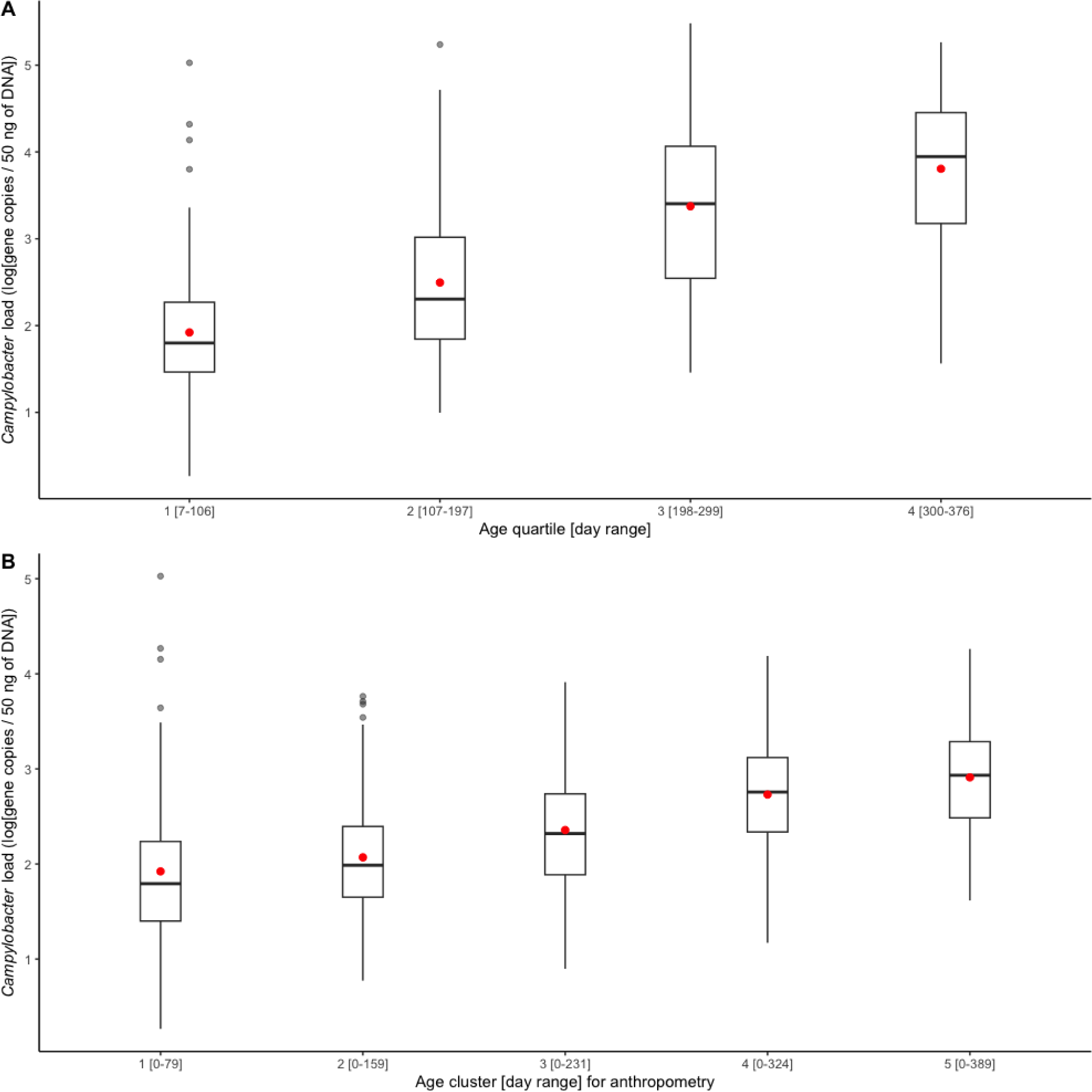
Boxplots of average *Campylobacter* load (A) by age quartile and (B) from birth to each length-for-age cluster. A red dot indicates the mean of each set.

The mean LAZ decreased from −0.45 (SD = 1.08) in age group 1 (mean age = 94 days) to −2.06 (SD = 0.93) in age group 5 (mean age = 428 days), see Figure 2 (A). Meanwhile, the prevalence of stunting increased steadily from 3% (95% CI: 1%-11%) to 51% (95% CI: 40%-61%) in the same time frame, see Figure 2 (B). EED measurements were conducted at an average age of 418 (SD = 35) days, and the prevalence of EED was 56% (95% CI: 46%-0.65%), of which 25% (95% CI: 17%-34%) were severe EED. Current diarrhea and fever prevalence rates were higher in the later age quartiles than in the first (Figure 3). Generally, the levels or prevalence of many time-varying determinants increased as the infants aged (Figure 4).

**Figure 2.**
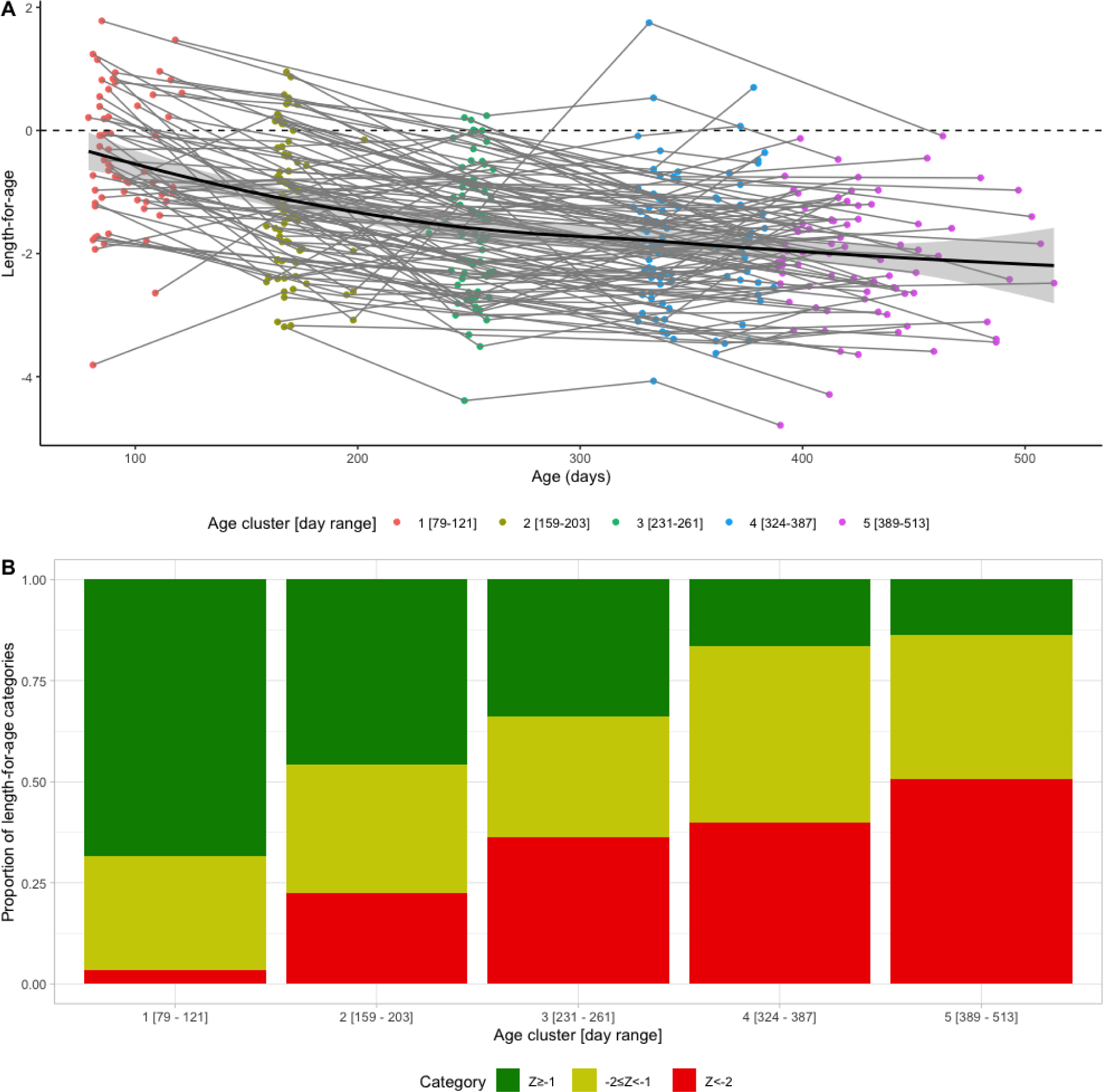
Longitudinal profiles of length-for-age (LAZ) scores (A) LAZ as a function of age for individual infants (thin lines), population average (solid line) and 95% confidence interval (shaded band). (B) LAZ categories by age cluster. Green: not stunted (LAZ ≥ −1), yellow: at risk of stunting (−2 ≤ LAZ < −1), red: stunted (LAZ< −2).

**Figure 3.**
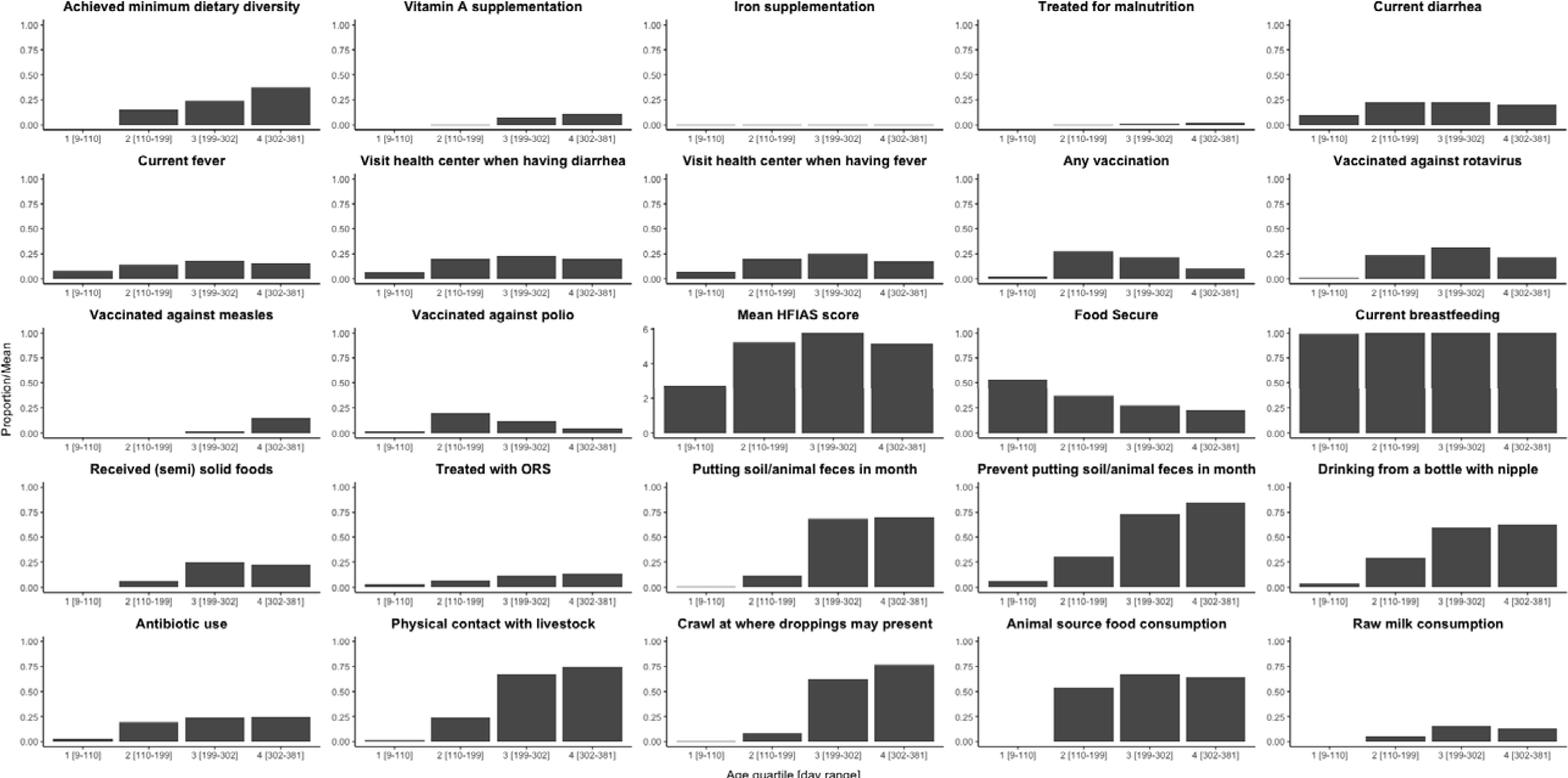
Proportions/averages of putative determinants by age quartile.

**Figure 4.**
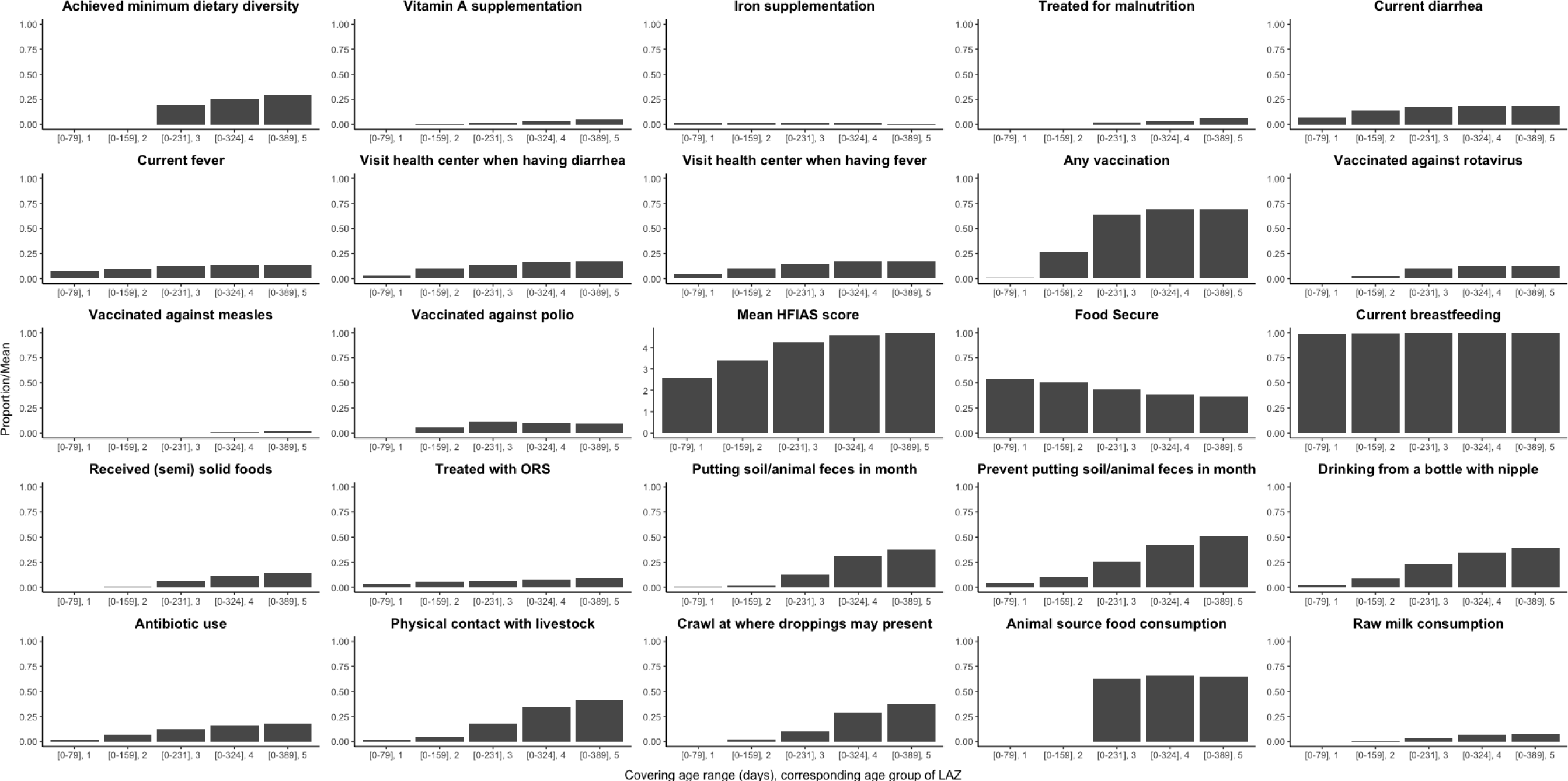
Average proportions/score of putative determinants from birth to each age cluster of visits for length-for-age Z score (LAZ).

However, we observed consistently low proportions of iron supplementation and treatment for malnutrition, a consistently high proportion of breastfeeding, and the highest proportions of vaccination (including polio and rotavirus vaccines) in the second age quartile. The introduction of complementary feeding was reported at a mean age of 187 days (range 46-319). Based on the HFIAS score, the proportion of the study population that was food secure decreased as infants aged.

### Regression analysis

#### Campylobacter

Variable screening results of the individual- and household-level determinants (both with and without adjustment for confounders) are summarized in Supplementary Table 1. Determinants with adjusted p values < 0.2 in the screening analyses were included in multivariable analyses. The final models for determinants of the outcomes of *Campylobacter* infection, at different time scales, is presented in Table 2. A lower average of *Campylobacter* load over the whole follow-up period was significantly associated with mothers’ hand washing with soap and a higher sheep nighttime risk score were, while a higher chicken daytime risk score and not disposing of infant stool outside the home were associated with a higher *Campylobacter* load. In the first month after birth (day range: [7–39]), an increasing *Campylobacter* load was strongly associated with prelacteal feeding and infant sex did not modify the effect for this association. The longitudinal model over age quartiles found that an increasing *Campylobacter* load was associated with complementary feeding, consumption of raw milk, and food insecurity.

**Table 2.**
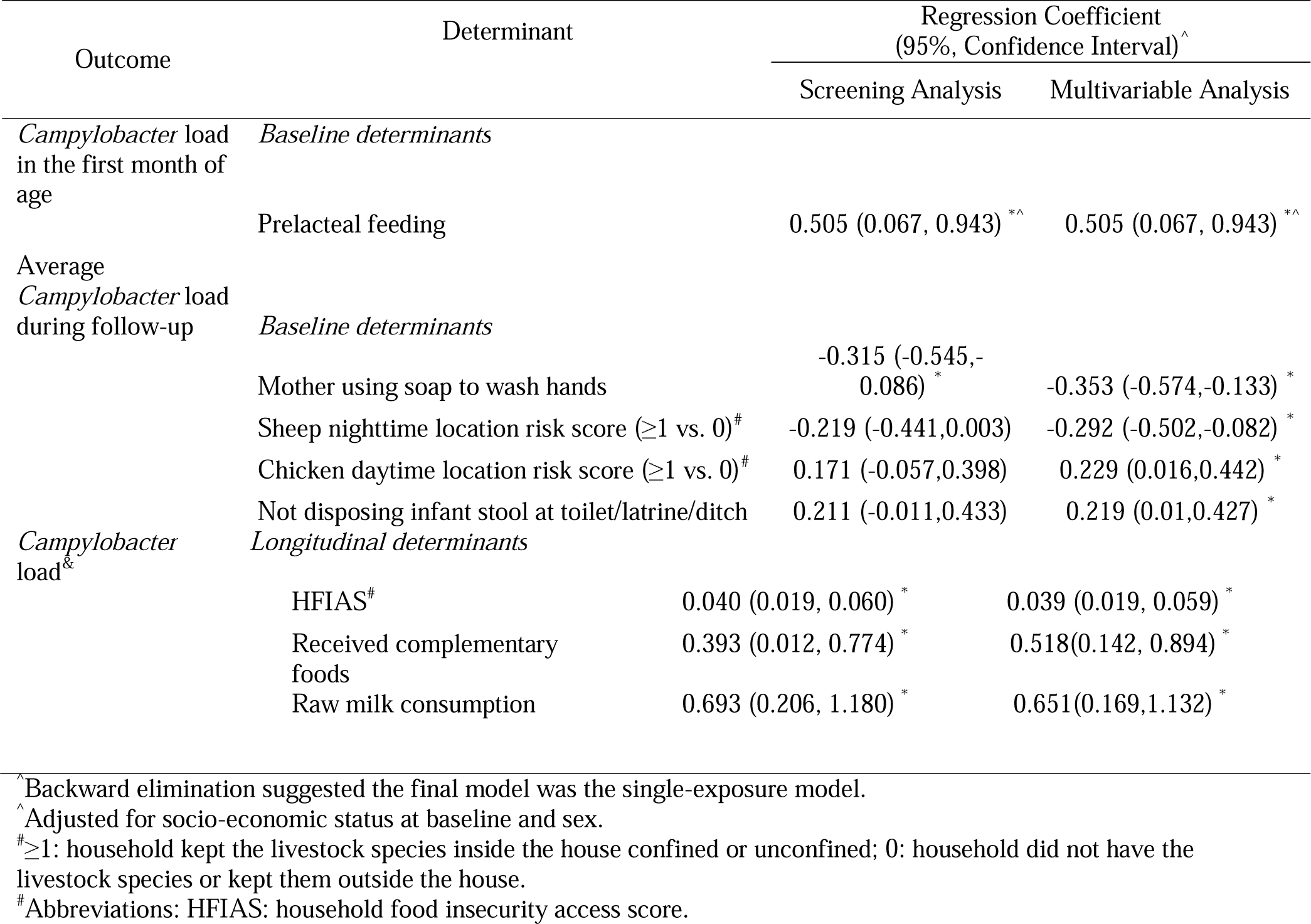
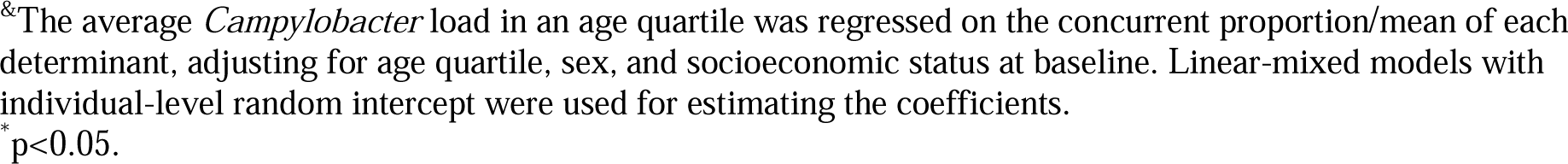
Associations between *Campylobacter* load and determinants.

| Outcome | Determinant | Regression Coefficient<br>(95%, Confidence Interval)<sup>^</sup> |  |
| --- | --- | --- | --- |
|  |  | Screening Analysis | Multivariable Analysis |
| *Campylobacter* load in the first month of age | *Baseline determinants* |  |  |
|  | Prelacteal feeding | 0.505 (0.067, 0.943) <sup>*^</sup> | 0.505 (0.067, 0.943) <sup>*^</sup> |
| Average *Campylobacter* load during follow-up | *Baseline determinants* |  |  |
|  | Mother using soap to wash hands | -0.315 (-0.545,-0.086) <sup>*</sup> | -0.353 (-0.574,-0.133) <sup>*</sup> |
| | Sheep nighttime location risk score ($\geq 1$ vs. 0)<sup>#</sup> | -0.219 (-0.441,0.003) | -0.292 (-0.502,-0.082) <sup>*</sup> |
| | Chicken daytime location risk score ($\geq 1$ vs. 0)<sup>#</sup> | 0.171 (-0.057,0.398) | 0.229 (0.016,0.442) <sup>*</sup> |
|  | Not disposing infant stool at toilet/latrine/ditch | 0.211 (-0.011,0.433) | 0.219 (0.01,0.427) <sup>*</sup> |
| *Campylobacter* load<sup>&</sup> | *Longitudinal determinants* |  |  |
|  | HFIAS<sup>#</sup> | 0.040 (0.019, 0.060) <sup>*</sup> | 0.039 (0.019, 0.059) <sup>*</sup> |
|  | Received complementary foods | 0.393 (0.012, 0.774) <sup>*</sup> | 0.518(0.142, 0.894) <sup>*</sup> |
|  | Raw milk consumption | 0.693 (0.206, 1.180) <sup>*</sup> | 0.651(0.169,1.132) <sup>*</sup> |

<sup>^</sup>Backward elimination suggested the final model was the single-exposure model.
<sup>^</sup>Adjusted for socio-economic status at baseline and sex.
<sup>#</sup> $\geq 1$ : household kept the livestock species inside the house confined or unconfined; 0: household did not have the livestock species or kept them outside the house.
<sup>#</sup>Abbreviations: HFIAS: household food insecurity access score.
<sup>&</sup>The average *Campylobacter* load in an age quartile was regressed on the concurrent proportion/mean of each determinant, adjusting for age quartile, sex, and socioeconomic status at baseline. Linear-mixed models with individual-level random intercept were used for estimating the coefficients. \*p<0.05.

Diarrhea and fever frequency were associated with a higher *Campylobacter* load, although the latter was not statistically significant after adjustment for confounders (Table 3).

**Table 3.** Associations between enteric disease symptoms and *Campylobacter* load.

| Outcome | Determinant | Regression Coefficient (95%, Confidence Interval)<sup>^</sup> |  |
| --- | --- | --- | --- |
|  |  | Unadjusted Analysis | Adjusted Analysis |
| Fever | *Campylobacter* load | 0.034 (0.014,0.054) * | 0.025 (-0.002,0.052) |
| Diarrhea |  | 0.040 (0.018,0.062) * | 0.041 (0.012,0.071) * |

<sup>^</sup>Using a linear mixed model with individual-level random intercept, the proportion of each symptom's occurrence was regressed as the outcome on the concurrent average *Campylobacter* load using crude and adjusted models (adjusting for age quartile, sex, and socioeconomic status at baseline). \*p<0.05.

#### EED

We found no significant associations between EED or individual indicators of gut function with *Campylobacter* load (Table 4).

**Table 4.** Associations between gut function indicators and *Campylobacter* load.

| Outcome | Determinant | Unadjusted odds ratio<br>(95% confidence interval) | Adjusted odds ratio<br>(95% confidence interval)<sup>^</sup> |
| --- | --- | --- | --- |
| Environmental enteric dysfunction | *Campylobacter* load | 1.136 (0.572,2.257) | 1.127 (0.557,2.283) |
| Fecal myeloperoxidase |  | 0.802 (0.404,1.593) | 0.736 (0.355,1.527) |
| Lactulose percentage |  | 0.649 (0.326,1.291) | 0.635 (0.315,1.282) |

<sup>^</sup>Adjusted for sex and socioeconomic status (indicated by a binary variable of household asset) at baseline, and age at EED sampling.

An increased odds of EED was associated with increased infant age at sampling and mother’s handwashing before infant feeding. A lower odds of EED was associated with increasing number of sheep in the household, mother washing her hands after field work, and being food secure (Table 5). A median level (0<proportion<0.15) of Vitamin A supplementation increased the odds of EED compared to no supplementation at all (proportion=0).

**Table 5.** Associations between Environmental Enteric Dysfunction and determinants.

| Outcome | Determinant | Odds ratio (95% CI)<sup>^</sup> |  |
| --- | --- | --- | --- |
|  |  | Screening Analysis | Multivariate Analysis |
| EED | *Baseline determinants* |  |  |
|  | Number of sheep (per head) | 0.81 (0.626,1.049) | 0.715 (0.532,0.960) * |
|  | Mother handwashing before feeding infant | 3.313 (0.998,10.994) | 5.963 (1.24,28.669) * |
|  | Mother handwashing after field work | 0.284 (0.082,0.983) | 0.179 (0.037,0.858) * |
|  | Achieved MDD<sup>#</sup> | 5.441 (0.929,31.856) | 10.904 (0.890,133.569) |
|  | *Longitudinal determinants<sup>§</sup>* |  |  |
|  | Proportion of Vitamin A supplementation<sup>^</sup> |  |  |
|  | ≥ 0.15 | 2.161 (0.460,10.158) | 0.52 (0.067,4.035) |
|  | > 0 & < 0.15 | 3.815 (1.378,10.563) * | 5.62 (1.529,20.651) * |
|  | Proportion being Food Secure | 0.192 (0.027,1.371) | 0.028 (0.002,0.502) * |
|  | Age (days) when an EED sample was taken<sup>#</sup> | 1.005 (0.993,1.018) | 1.024 (1.005,1.043) * |

<sup>#</sup>Abbreviations: EED: environmental enteric dysfunction, MDD: minimum dietary diversity, CI: confidence interval.
<sup>§</sup>We regressed EED on the overall proportion of each individual for longitudinal determinants (i.e., achieved MDD, proportion of Vitamin A supplementation, proportion being Food Secure).
<sup>^</sup>For Multivariable analysis, we used likelihood-ratio test (LRT) to select the final model and reported Wald-typed 95% CIs. Note that “Achieved MDD” had a P value of 0.049 by LRT in the final model.
<sup>^</sup>We categorized the proportion into three categories, ≥ 0.15, > 0 & < 0.15, and =0 (for the reference group) to strengthen the robustness of 95% CI of this variable.
\*p<0.05.

#### LAZ

Despite initial findings in a sex- and SES-adjusted linear-mixed model suggesting that the average *Campylobacter* load had a significant strong effect on increasing growth faltering (regression coefficient: 0.54, P value: <0.001), this effect was no longer significant and had a drastic reduction in effect size after adjusting for age (regression coefficient: 0.01, P value: 0.897) (Supplementary table 1). There were no significant associations between the change in the LAZ score from baseline to endline and indicators of gut function (Table 6).

**Table 6.**
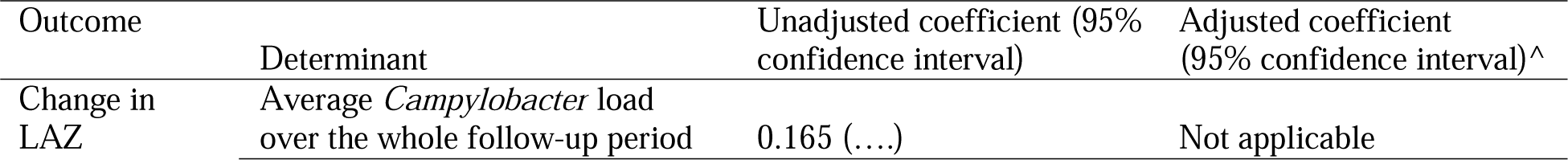
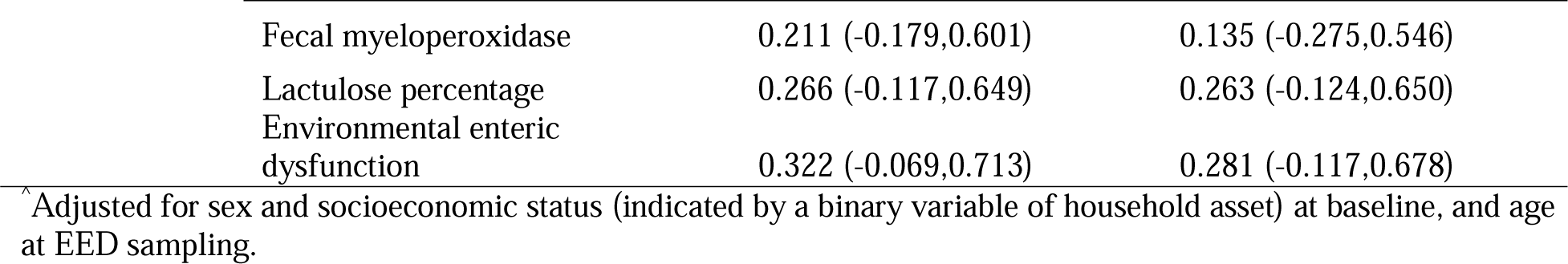
Associations between change in LAZ from baseline to endline, *Campylobacter* load and gut function indicators.

| Outcome | Determinant | Unadjusted coefficient (95% confidence interval) | Adjusted coefficient (95% confidence interval)<sup>^</sup> |
| --- | --- | --- | --- |
| Change in LAZ | Average *Campylobacter* load over the whole follow-up period | 0.165 (....) | Not applicable |

|  |  |  |
| --- | --- | --- |
| Fecal myeloperoxidase | 0.211 (-0.179,0.601) | 0.135 (-0.275,0.546) |
| Lactulose percentage | 0.266 (-0.117,0.649) | 0.263 (-0.124,0.650) |
| Environmental enteric dysfunction | 0.322 (-0.069,0.713) | 0.281 (-0.117,0.678) |

<sup>^</sup>Adjusted for sex and socioeconomic status (indicated by a binary variable of household asset) at baseline, and age at EED sampling.

Variable screening results of the individual- and household-level determinants for the change in LAZ from baseline to endline (both with and without adjustment for confounders) are summarized in Supplementary Table 1. Determinants with adjusted p values < 0.2 in the screening analyses were included in multivariable analyses. The final model for baseline determinants of LAZ is presented in Table 8. Note: Change in LAZ was calculated as (first available LAZ – last available LAZ). Hence, positive regression coefficients indicate a *greater decrease* in LAZ and thus are associated with *poorer* nutrition outcomes. Longitudinally, a greater decrease in LAZ (poorer nutrition outcome) was associated with mothers’ hand washing after cleaning infants post-defecation, and infants being female, while mother having attended school, keeping chickens inside the household during the daytime (i.e., chicken daytime location risk score ≥1), and an infant being underweight at enrollment were associated with a smaller decrease in LAZ (better nutrition outcome). Longitudinally, a decrease in LAZ was associated with more household food insecurity (Table 9).

**Table 8.**
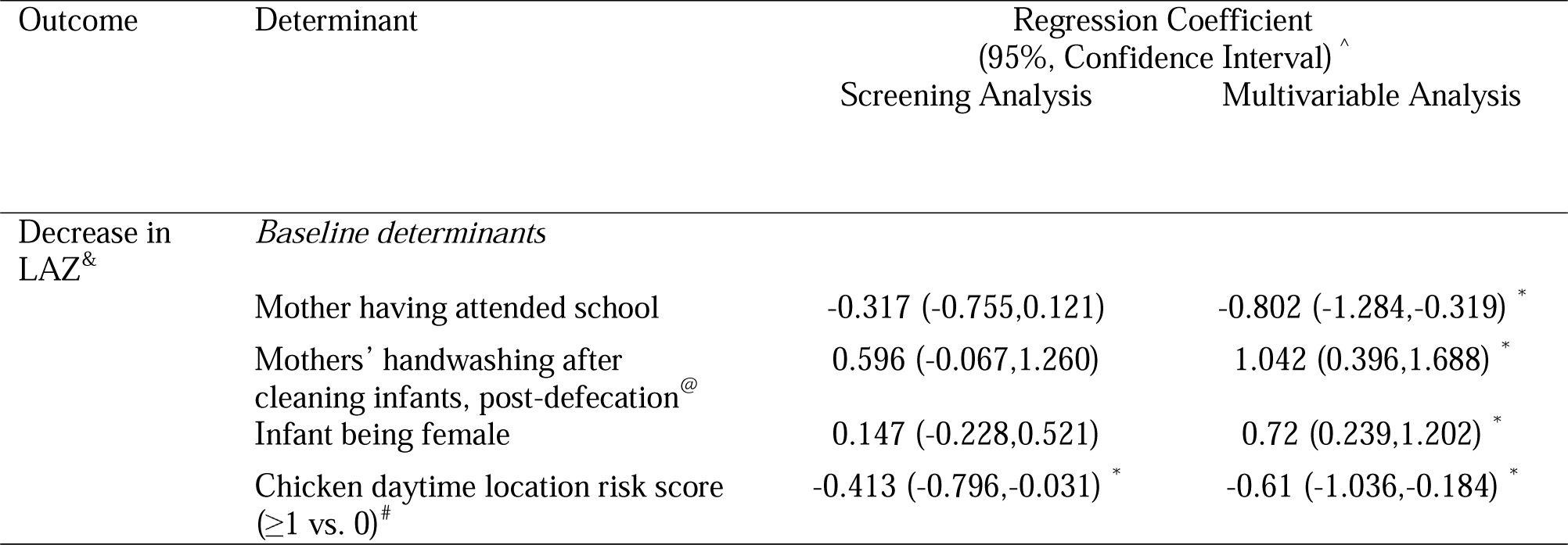
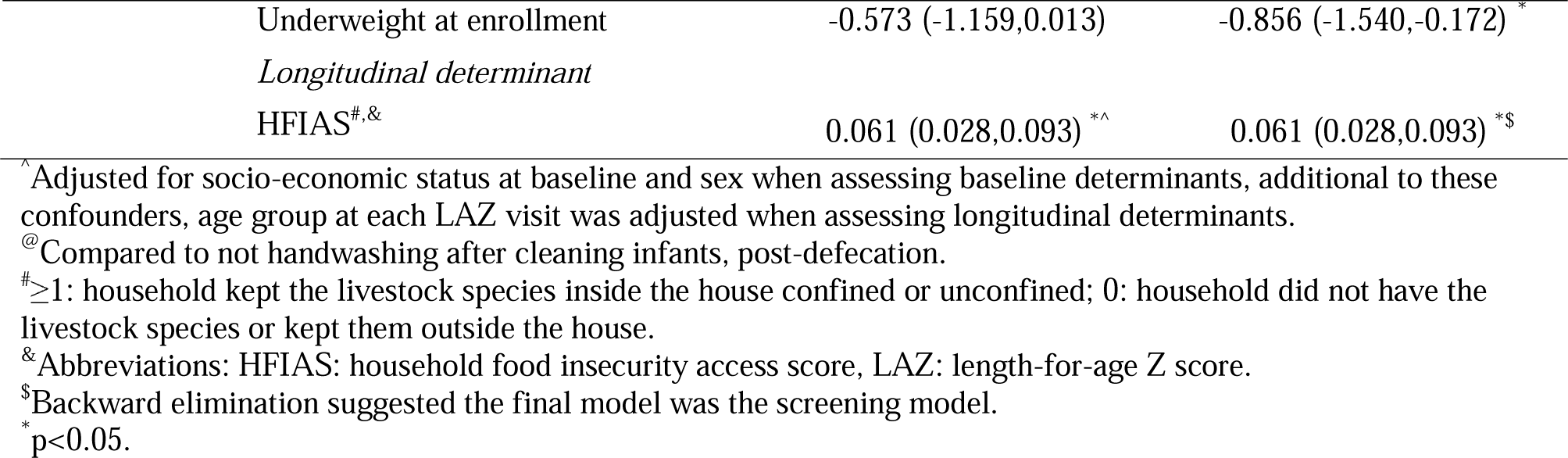
Associations between household determinants and decrease in LAZ.

| Outcome | Determinant | Regression Coefficient<br>(95%, Confidence Interval) <sup>^</sup> |  |
| --- | --- | --- | --- |
|  |  | Screening Analysis | Multivariable Analysis |
| Decrease in LAZ<sup>&</sup> | *Baseline determinants* |  |  |
|  | Mother having attended school | -0.317 (-0.755,0.121) | -0.802 (-1.284,-0.319) * |
|  | Mothers' handwashing after cleaning infants, post-defecation<sup>@</sup> | 0.596 (-0.067,1.260) | 1.042 (0.396,1.688) * |
|  | Infant being female | 0.147 (-0.228,0.521) | 0.72 (0.239,1.202) * |
| | Chicken daytime location risk score ($\geq 1$ vs. 0)<sup>#</sup> | -0.413 (-0.796,-0.031) * | -0.61 (-1.036,-0.184) * |

|  |  |  |
| --- | --- | --- |
| Underweight at enrollment | -0.573 (-1.159,0.013) | -0.856 (-1.540,-0.172) * |
| Longitudinal determinant |  |  |
| HFIAS<sup>#,&</sup> | 0.061 (0.028,0.093) <sup>*,^</sup> | 0.061 (0.028,0.093) <sup>*,\$</sup> |

<sup>^</sup>Adjusted for socio-economic status at baseline and sex when assessing baseline determinants, additional to these confounders, age group at each LAZ visit was adjusted when assessing longitudinal determinants.
<sup>@</sup>Compared to not handwashing after cleaning infants, post-defecation.
<sup>#</sup><sub>≥1</sub>: household kept the livestock species inside the house confined or unconfined; 0: household did not have the livestock species or kept them outside the house.
<sup>&</sup>Abbreviations: HFIAS: household food insecurity access score, LAZ: length-for-age Z score.
<sup>\$</sup>Backward elimination suggested the final model was the screening model.
<sup>\*</sup>p<0.05.

## Discussion

In this prospective birth cohort, infants experienced linear growth faltering during the first year of life. More than half of the infants were stunted (LAZ <-2) by endline (day range 389-513), up from 3% in months 3-4 (day range 79-121). More than half (56%; 95% CI: 46%-0.65%) of infants had EED at the end of the longitudinal study, in which 25% (95% CI: 17%-34%) had severe EED. More than 30% of infants were colonized with *Campylobacter* in the first month of life, and the prevalence increased continuously, reaching a plateau of nearly 90% between months 8 and 9 [39]. Despite the high occurrence of *Campylobacter* infection, EED, and stunting in this population, no association was found between *Campylobacter* load and EED, EED and stunting, or *Campylobacter* load and stunting.

Several determinants for *Campylobacter* load, EED, and LAZ were identified. Reported practice of soap use by mothers during handwashing was significantly associated with a lower *Campylobacter* load, as well as was the sheep nighttime location risk score, indicating that increased contact with sheep at night is associated with a lower *Campylobacter* load. Conversely, a higher chicken daytime location risk score, as well as not disposing of infant stool outside the home, were both associated with an increased *Campylobacter* load. Prelacteal feeding contributed to an increase in *Campylobacter* load in the first month of life. When longitudinal data were analyzed, risk factors associated with increased *Campylobacter* load included complementary feeding, consumption of raw milk, and food insecurity. Among time-varying determinants, a higher *Campylobacter* load was associated with more frequent concurrent diarrhea. Determinants for EED and LAZ were also identified. A greater probability of EED was associated with older age of the infant at sampling, and mothers reporting handwashing prior to infant feeding. A lower probability of EED was associated with household food security and the mother reporting handwashing after field work. Larger decrease in LAZ of infants, a poorer health outcome, was associated with being female and the mother reporting handwashing after infant cleaning post-defecation. Mother having attended school, the infant being underweight at enrollment (age range 4-27 days), and keeping chickens inside without confinement during daytime were associated with *less* decrease of LAZ, a better growth outcome. Among time varying determinants, only food insecurity was associated with higher risks of linear growth faltering.

Similar to the MAL-ED study, we observed a sharp increase in *Campylobacter* prevalence in the first year of life; however, nearly all infants were colonized with *Campylobacter* by month 13, which was higher than the prevalence rates observed at that time across MAL-ED sites. Such prevalence increase may be driven by a high force of infection (FOI) of and/or a weakened clearance against *Campylobacter* [40]. The observed difference between studies may also be due to the difference in sensitivity and specificity of assays used for testing *Campylobacter*, thus caution should be taken when attempting to compare pertinent results (e.g., prevalence) between studies. A high FOI is determined by frequent environmental exposures, as observed in our setting. Stunting-associated immunosuppressive effects might weaken clearance capacity. This also implies at least the potential for reverse causality (namely, stunting increases infection) as depicted in the “vicious cycles of diseases of poverty” [41].

Our findings reveal that food insecurity was associated with both higher *Campylobacter* load and growth faltering which is in line with the role that this determinant plays as an underlying cause in the modified UNICEF framework for undernutrition [9]. The significant contribution of raw milk consumption to increasing *Campylobacter* load is consistent with findings from our formative research, where we found a significant association between animal-sourced foods consumption (primarily consisting of raw milk) and increasing *Campylobacter* detection in young children [9]. Our results indicate that complementary feeding may be a source of enteric infection by *Campylobacter*. The risk of complementary feeding may be reduced by optimizing or improving existing handwashing behaviors (such as mother’s handwashing with soap), as handwashing (particularly with soap) has been associated with the reduction of microbial contamination in complementary foods [42].

Being in close proximity to sheep at night was found to be associated with lower *Campylobacter* load, where closer proximity to chicken throughout the day was found to be associated with higher *Campylobacter* load. These contradictory results for risks associated with animals suggest the relationships between animal ownership and *Campylobacter* colonization are highly complex and may be confounded by the lack of adjustment for *Campylobacter* species-level infection. We are undertaking an MLST-based attribution study that aims to tease out the relationships between livestock and Campylobacter infection at the species level.

The practice of prelacteal feeding is common in LMIC despite its potential risks [43]. A 2018 meta-analysis suggested the prevalence of this practice in Ethiopia was around 25% [44]. A more recent study of children under 2 years conducted near our study area suggested that nearly half of the population received a prelacteal feeding (a cultural practice in the region), which was associated with a lack of knowledge about the risks related to the practice [45]. In comparison, the prevalence of prelacteal feeding was even higher in our cohort (67%). Given that the practice is the sole significant feeding practice associated with an increased *Campylobacter* load in the first month of life, and this was independent of sex, future research may be needed to better understand what drives the practice, the cultural meanings assigned to the practice, and possible interventions to strengthen mothers’ knowledge of its risks.

Comparing the common putative determinants in this study with our cross-sectional study [24], most of them were qualitatively similar. However, overall, we observed fewer livestock holdings in the longitudinal study, which may be related to the timing of our studies. The cross-sectional work was conducted in 2018, while the data presented here were collected after the onsets of both the COVID-19 pandemic and the civil war in Tigray, Ethiopia. Both disruptions and their consequential instability were likely to have caused financial distress, which may have triggered the selling of livestock for income.

The prevalence of child EED in our study population (56%) was similar to findings from the formative research (50%, 95% CI: 40–60%), severe EED was higher (25%) but not significantly different from the formative research (17%, 95% CI: 11–26%) [24]. The lower odds of EED found associated with food security and mothers handwashing after field work both align and are consistent with broader understandings of EED and its relationship hygiene and sanitation and other socioeconomic factors [12]. That an increased odds of EED was associated mother’s handwashing before feeding the infant and supplementation of Vitamin A are additional counter-intuitive findings that merit exploration but could also reflect residual confounding.

Despite having a lower prevalence of stunting at the first visit around 3-4 months old, the trend of LAZ in the study population is similar to that which was observed in Haydom, Tanzania (TZH) in the MAL-ED study [46], which was also rural and comprised of smallholder households who relied heavily on corn production as a cash crop [47]. Our study population engaged in khat production, another cash crop, as their primary livelihood, the implications of which have been explored elsewhere, with no significant impact on infant nutrition having emerged [48,49]. However, the comparability of the LAZ trends between our site and TZH may reflect a typical trajectory of infant nutrition in rural smallholder households where cash crop agriculture dominates [50].

Previously published literature using enrollment weight (collected ≤ 17 days of birth) as a surrogate measure for birth weight, found lower weight-for-age at this time increased the odds of being stunted [46]. In contrast, being underweight within 27 days of birth in our cohort was associated with *less* growth faltering. This finding might suggest more attentive care or feeding practices of infants with visible clinical signs of being underweight, resulting in some catch-up growth. Contrary to much literature that finds being male to be a risk factor for stunting [51], female infants had more growth faltering in our setting. Existing literature indicates that any effect of biological sex on linear growth may be modified by its complex interactions with social factors [52]. Our study was conducted in a traditional, rural population of Sunni Muslims, where gender norms in society may be favorable for males [53], including infants. Given increasing evidence surrounding the importance of gender inequality as a predictor of child stunting, cultural norms may explain the increased risk of growth faltering among girls [54]. A similar finding was observed in Bangladesh, a Muslim-majority country, where female children had lower LAZ [55]. The associations between linear growth faltering and the determinants of chicken daytime risk score and the mother reporting handwashing following cleaning an infant after defecation, in opposite directions, are counterintuitive. The following hypothesized scenarios could explain these counterintuitive associations: First, more chickens might indicate higher access to a protein source or more female-controlled revenue, which might outweigh the risk for infant growth. There is increasing evidence that in this study setting women’s empowerment is strongly associated with infant growth outcomes [48,49]. Second, mothers who report handwashing after cleaning infants post-defecation may be more likely to clean their own infants as opposed to having an older child or another person in the house clean the infant; or mother’s hands could be re-contaminated in between the handwashing after cleaning the infant and other infant care practices, thereby exposing the infant to *Campylobacter* and increased growth faltering. Importantly, a large body of evidence supports the understanding that infant growth, and stunting in particular, is multi-causal. Given the small sample size of this analysis, complex models with numerous interacting determinants are not possible. Thus, some of these associations may be spurious. Scenarios like those presented above could not be tested in this study through available instruments. However additional investigation, using parallel observational data collection methods on the same population (see forthcoming literature from the EXCAM study), aim to better understand some of the counter-intuitive findings.

Built on the formative research, the data from this longitudinal study offers a more thorough understanding of the socio-demographic and exposure landscapes in the Haramaya woreda and similar smallholder settings. The repeated-measures short survey enabled our research team to study the health outcomes at different time scales and allowed a more accurate characterization of the timings related to Infant and Young Child Feeding (IYCF).

This study has limitations. This study was powered for prevalence estimation of the *Campylobacter* genus [25], limiting our ability to evaluate statistical significance in the regression analyses such as the key association between *Campylobacter* infection and growth faltering. This was further constrained by an increased level of missing data due at least in part to our field work being disrupted by the global pandemic and civil unrest [25]. Despite the adjustment for common confounders (sex, age, SES) in the regression analyses, as previously mentioned, potential residual confounding, a common problem in survey-based observational studies, could bias the associations of interest. In addition, common survey biases might exist in the survey data. For example, recall bias could exist in the monthly complementary feeding status; and responses to questions about soap use, in our setting where soap use was rare, could be subject to social desirability bias. With a relatively small study population size, the screening of a large set of variables in regression analyses may subject to type I error from multiple comparisons.

In summary, this work uncovered an increasing *Campylobacter* load in the first year of life, a high prevalence of EED by 12-13 months of age, and a concurrent increasing level of growth faltering, none of which were significantly associated with the other, but all three of which were associated with food insecurity. Despite a growing consideration of exposure to livestock feces in both this research and current interventions to improve child growth [22], our findings around livestock risk factors were inconclusive. We found that even though livestock may be the reservoir of *Campylobacter* infections in infants, these relationships are highly complex and most determinants of the infection in this study were more proximal to the infant, i.e., related to improper IYCF practices and WaSH in our setting. This signifies a need to strengthen both IYCF practices and infant food safety and hygiene to reduce (cross-) contamination at the point of exposure and, ultimately, reduce the risk of infection and improve linear growth. Future research should aim to replicate findings in diverse populations to assess the generalizability of results and identify context-specific factors influencing *Campylobacter* transmission and health outcomes.

## Data Availability

Deidentified individual participant data will be made available through Dataverse (https://dataverse.org/) after December 31, 2024.

## Supplementary material

Supplementary Table 1. Determinants of outcomes of *Campylobacter* load and/or decrease in length-for-age Z score.

Supplementary Table 2. Determinants of environmental enteric dysfunction.

## Acknowledgments

This work is a result of the CAGED Research Team. Beyond those named authors, other CAGED Research Team members include: Cyrus Saleem, Efrah Ali Yusuf, Getnet Yimer, Kunuza Adem Umer, Karah Mechlowitz, Mawardi M. Dawid, Mahammad Mahammad Usmail, Wondwossen A. Gebreyes, Yenenesh Demisie Weldesenbet, Zelalem Hailu Mekuria. This study would not have been possible without cooperation of study communities and local administration of the study kebeles. We would like to express our appreciation to the study participants, their households, the Community Advisory Board, and all who supported the study directly or otherwise.

We thank Sridevi Devaraj PhD Texas Children’s Hospital, Baylor College of Medicine, Houston, TX for analysis of lactulose in urine.

Research reported in this publication was supported by the University of Florida Clinical and Translational Science Institute, which was supported in part by the NIH National Center for Advancing Translational Sciences under award number UL1TR001427. The content is solely the responsibility of the authors and does not necessarily represent the official views of the National Institutes of Health.

## Funding

This project is funded by the United States Agency for International Development Bureau for Food Security under Agreement #AID-OAA-L-15-00003 as part of Feed the Future Innovation Lab for Livestock Systems, and by the Bill & Melinda Gates Foundation OPP#1175487. Under the grant conditions of the Foundation, a Creative Commons Attribution 4.0 Generic License has already been assigned to the Author Accepted Manuscript version that might arise from this submission. Any opinions, findings, conclusions, or recommendations expressed here are those of the authors alone.

### Declarations

#### Ethics approval and consent to participate

Ethical approval was obtained from the University of Florida Internal Review Board (IRB201903141); the Haramaya University Institutional Health Research Ethics Committee (COHMS/1010/3796/20) and the Ethiopia National Research Ethics Review Committee (SM/14.1/1059/20). Written informed consent was obtained from all participating households (husband and wife) using a form in the local language (Afan Oromo).

#### Consent for publication

Not applicable.

